# Should women have lower blood pressure targets than men? Sex differences in blood pressure and cardiovascular disease in the UK Biobank

**DOI:** 10.1101/2024.09.03.24313046

**Authors:** Rebecca K Kelly, Katie Harris, Cheryl Carcel, Paul Muntner, Mark Woodward

## Abstract

**Background:** Recent studies show that the risk of cardiovascular disease (CVD) increases from a lower nadir of systolic blood pressure (SBP) in women than men, and increases thereafter at a greater rate. This has led to a suggestion that sex-based SBP thresholds are required. We aimed to investigate sex differences in the associations of SBP and incident atherosclerotic CVD in a large prospective cohort.

**Methods:** 420,649 UK Biobank participants with no prior history of CVD were included. Age-adjusted sex-specific risks, relative risks (RRs) and risk differences (RDs) relating SBP to CVD were estimated using Poisson and Cox regression.

**Results:** Over 13.2 years of follow-up there were 28,628 CVD events. CVD risks across BP levels showed a “J-shape”, and were higher in men than women at all BP levels. The lowest risks were at SBP 100-<105 mmHg (events per 10,000 person-years [95%CI]: 15.6 [11.8-23.1]) and 110-<115 (47.2 [41.8-53.0]) among women and men, respectively. Compared with SBP 100-<110, sex-specific RRs at above 120 were higher in women than men, but RDs were higher in men than women at all levels of SBP. Furthermore, compared to men at 100-<110 (i.e. the men with least risk), risks in women were lower at all levels of SBP below 170.

**Conclusions:** CVD risk is lowest for women at a slightly lower SBP than men and RRs for CVD increase with SBP at a slightly steeper rate in women. However, both risks and RDs in women are never greater than in men. This evidence suggests that women should not have lower BP thresholds.

**Clinical Perspective:** *What is New?:*

- While risks are higher at lower BP levels in women than men when both sexes are considered independently, consideration of the risks and sex-combined analyses do not support treating women at lower BP levels than men.

*What are the Clinical Implications?:*

- In line with 2017 ACC/AHA guideline, antihypertensive treatment thresholds should be based on absolute risk calculated from sex-specific algorithms, rather than BP level in isolation.
- Both risks and relative risks must be taken into account to understand sex-differences in the relationship of BP-CVD and whether these differences are clinically meaningful.

## Introduction

Hypertension guidelines do not currently provide sex-based recommendations for initiating blood pressure (BP)-lowering therapy, citing limited evidence.^1–3^ However, a recent scientific statement from the European Society of Cardiology acknowledges that sex-specific thresholds for hypertension may be reasonable because the risk for cardiovascular disease (CVD) associated with higher BP starts at lower levels in women than men.^3^

Several observational studies have found that the relationship between BP and CVD differs by sex.^4–8^ These studies found higher relative risks (RRs) of CVD, compared to a low BP reference level, for women than men, as well as a lower nadir in the “J-shaped” association between BP and CVD in women than men.^4,8^ This has raised the question as to whether women should routinely be treated with antihypertensive medication at lower BP levels than men.^3,9^ However, none of these studies has reported this in the context of sex-specific risk differences (RDs), rather than RRs, or compared the RRs for women and men against a common reference group. In addition, several previous meta-analyses, have not found substantial sex difference in RRs for the relationship of BP with CVD events,^10–12^ or in the efficacy of BP lowering treatments.^13^

To address these uncertainties, we sought to use data from a large cohort of data, with more CVD events than has been previously analyzed, enabling precise measures within smaller ranges of BP than have previously been used. Furthermore, we examine whether the sex-differential association between BP and CVD differs by age, and the effect menopause has on the BP-CVD relationship in women.^14^

## Methods

### Subjects and study design

The UK Biobank is a prospective cohort study that includes approximately 500,000 men and women recruited between 2006 and 2010.^15,16^ Eligible adults aged between 40-69 years were invited to visit one of 22 centers for the baseline assessment, where detailed information on lifestyle and medical history was collected via a Touchscreen questionnaire and face-to-face interview, and physical measurements and biological samples were collected. Participants have been followed up for various health-related outcomes via linking to electronic health records. Further details, including the study protocol and data access permissions, are available online, and recruitment methods are described in detail elsewhere.^16^ All individuals provided informed written consent to participate and the study was approved by the National Information Governance Board for Health and Social care and the National Health Service North West Multicentre Research Ethics Committee (reference number 21/NW/0157). Bona fide researchers can apply to use the UK Biobank dataset by registering and applying at http://ukbiobank.ac.uk/register-apply/.

Participants were excluded from the present study if they withdrew consent (n=961), or had a history of CVD (self-reported or via linked electronic health records) prior to the baseline assessment, n=40,270), or did not have two automated measurements of systolic BP (SBP) and diastolic BP (DBP) at baseline after exclude excluding extreme measurements (n=41,437) (Figure S1).

### Measurement of blood pressure

SBP and DBP, in millimeters of mercury, were calculated as the mean of two measurements taken at least one minute apart, taken using an Omron HEM-7015IT digital BP monitor (Omron Healthcare). Hypertension was categorized using the American Heart Association (AHA) 2017 guidelines (no hypertension: SBP <130 mmHg and DBP <80 mmHg; and hypertension: SBP ≥130 mmHg or DBP ≥80 mmHg).^17^

### Outcomes

The primary outcome was incident atherosclerotic CVD, defined as a primary diagnosis of fatal or non-fatal coronary heart disease (CHD) or stroke. Secondary outcomes were incident CHD and stroke. Participant information on the date and cause of hospital admission(s) or death was obtained through linkage to hospital inpatient and death data.^18^ The present analyses were censored separately for incident CVD outcomes at the date of diagnosis or death, loss to follow-up, or administrative censoring for each country (31^st^ October 2022 for England, 31^st^ August 2022 for Scotland, 31^st^ May 2022 for Wales) whichever occurred first. CHD was defined by a primary diagnosis of incident (fatal or non-fatal) CHD (ICD-10 [international classification of diseases, 10th revision] codes I21-I25) or coronary revascularization (OPCS-4 [Classification of Interventions and Procedures, 4th revision] codes K49, K50, K75, K40-K46). Stroke was defined by a primary diagnosis of incident (fatal or non-fatal) stroke (ICD-10 codes I60-I61, I63-I64).

### Statistical analyses

Baseline characteristics for the overall population and women and men separately are presented as number (percentage) for categorical variables, or mean (standard deviation) or median (interquartile interval) for continuous variables as appropriate.

Sex-differences in the relationship of BP with CVD outcomes were explored using three approaches: (i) sex-specific risks and risk differences, (ii) sex-specific RRs, and (iii) sex-combined RRs. Poisson regression models were used to obtain age-adjusted rates and rate differences of incident CVD events (per 10,000 person-years), by sex and BP group.^19^ Cox proportional hazards regression models were used to estimate hazard ratios for CVD.^19^ For simplicity and ease of interpretation, Poisson-derived rates and Cox-derived hazards ratios are hereafter referred to as ‘risks’ and ‘relative risks’ (RRs), respectively. SBP and DBP were modelled using ordinal groups of 5 mm Hg, with reference groups for BP chosen based on previous literature.^4,8^ For sex-specific analyses, SBP 110-<115 mmHg and DBP 70-<75 mmHg in women and men were taken as the reference groups for each sex taken separately. For sex-combined analyses, men with SBP 100-<115 mmHg and men with DBP 70-<75 mmHg were taken as the common reference groups for both sexes. Then the sex-specific BP-CVD association was modelled continuously in increments of 10 mmHg for SBP and 5 mmHg for DBP, for both sexes. Finally, BP was modelled using the 2017 ACC/AHA BP guideline categories.^17^

All analyses were adjusted for age at recruitment (years), and multivariable models were additionally adjusted for the Townsend deprivation index (fifths from least to most affluent using national cut-off points, unknown), smoking status (never, former, light smokers [<15 cigarettes/d], medium smokers [15-<30 cigarettes/d], heavy smokers [≥30 cigarettes/d], smoker of unknown number of cigarettes, unknown), body mass index (BMI; kg/m^2^, unknown), diabetes (yes, no, unknown), antihypertensive medication use (yes, no, unknown), lipid-lowering medication use (yes, no, unknown), total cholesterol (mmol/L), and high-density lipoprotein cholesterol (HDL-C; mmol/L). Percentages of missing for each covariate were <7%, except for HDL-C (14.0%) (Table S1).

#### Subgroup analyses

To investigate whether sex differences in the associations of BP with CVD differed by age group (<50, 50-<55, 55-<60, and ≥60 years), we added a three-way interaction between sex, age group, and the BP exposure of interest to the models. To assess trend across age groups separately for men and women (P-trend) and in the three-way interaction (P-interaction) we used likelihood ratio tests (LRTs). Similarly, subgroup analyses were conducted by menopausal status (pre-menopausal, menopausal) in women, which was defined by self-report and other relevant factors (such as bilateral oophorectomy and use of hormone-replacement therapy).^20^

#### Sensitivity analyses

We conducted a sensitivity analysis restricting to participants not taking antihypertensive medication at baseline. Also, to assess the potential influence of regression dilution bias ^21^ we calculated sex-specific regression dilution ratios in a subsample of participants with BP measurements taken, on average, 10.6 years apart (n=51,375, 8.2% of the main study sample). RRs were corrected for regression dilution bias by multiplying the regression coefficients by the reciprocal of the regression dilution ratio.

We tested the proportional hazards assumption for our Cox models using Schoenfeld residuals, which showed no evidence of violation for exposures and covariates of interest in multivariable models for any outcome.

STATA version 18.0 (StataCorp LP, College Station, Texas) was used for data analyses and R 4.3.3 (R Core Team, Vienna, Austria) was used to create figures.

## Results

The baseline characteristics of 420,649 participants (56.0% women) without prior CVD included in the present analyses are shown in **Table 1**. At baseline, a lower proportion of women than men had hypertension, or taking antihypertensive or lipid lowering medication. Women had higher total cholesterol and HDL-C, and lower BMI. Among both women and men, participants with hypertension were older and had higher BMI, and a higher proportion were taking antihypertensive or lipid-lowering medication (Table S2). See Tables S3-S4 for baseline characteristics stratified by sex and SBP categories.

**Table 1.**
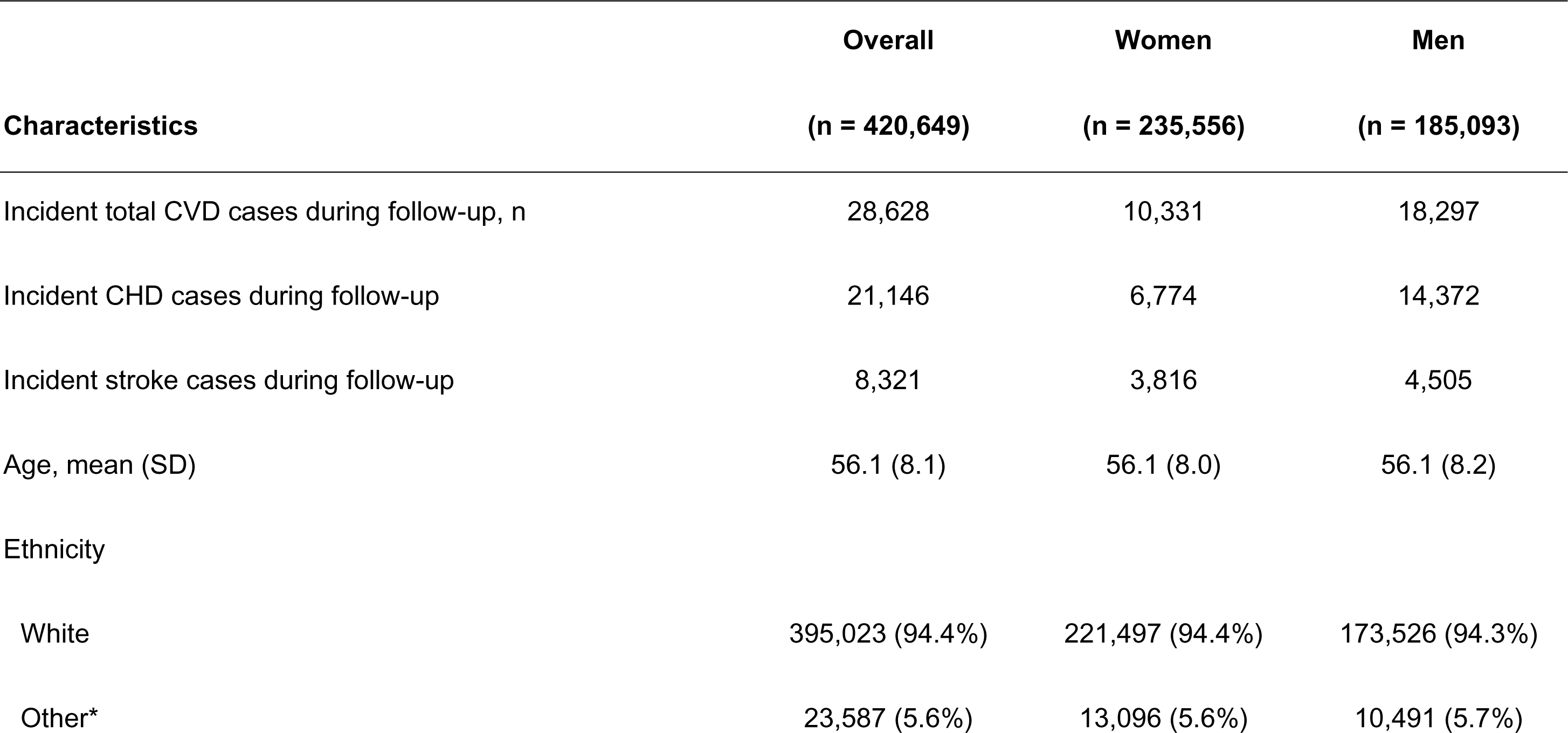
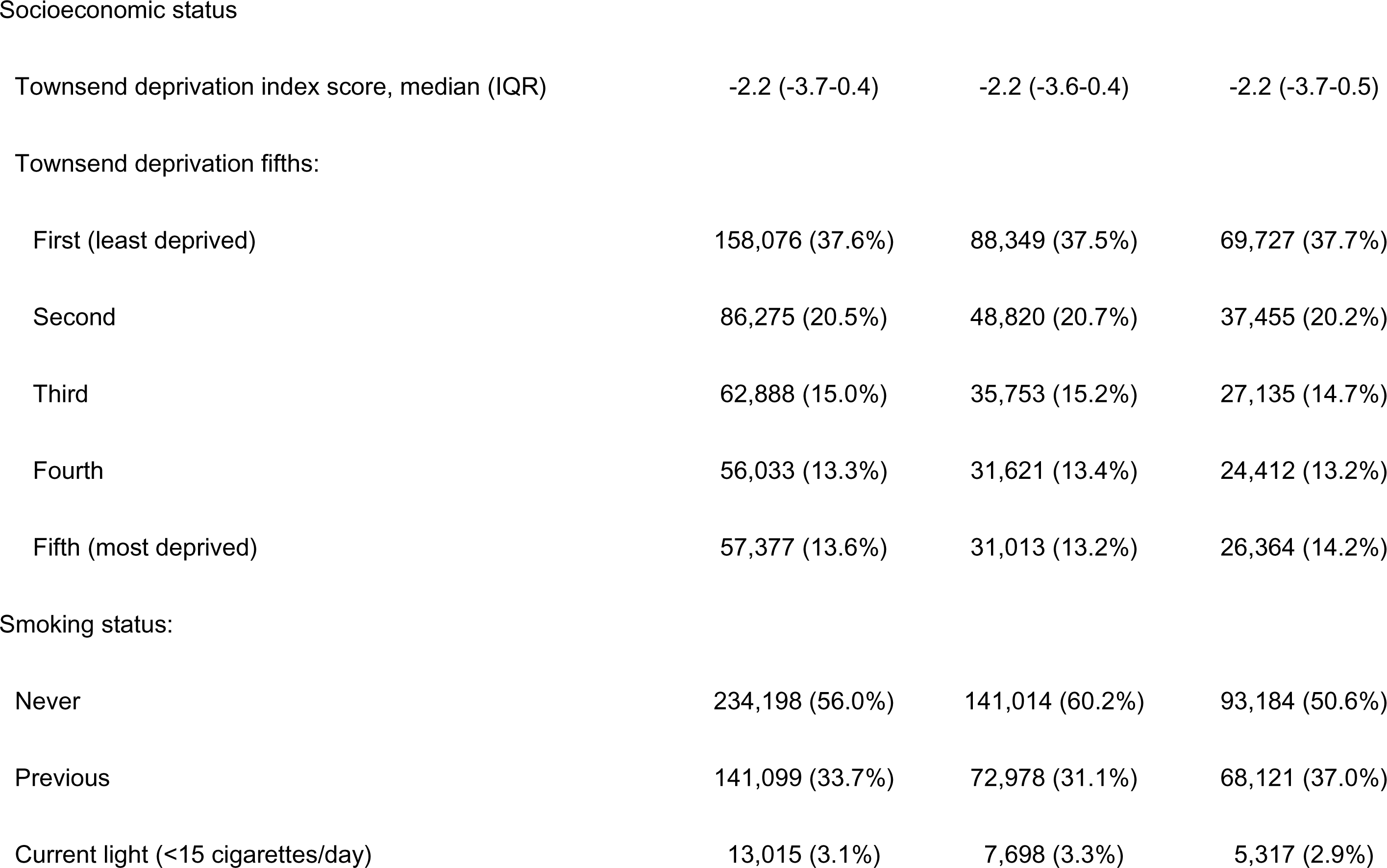
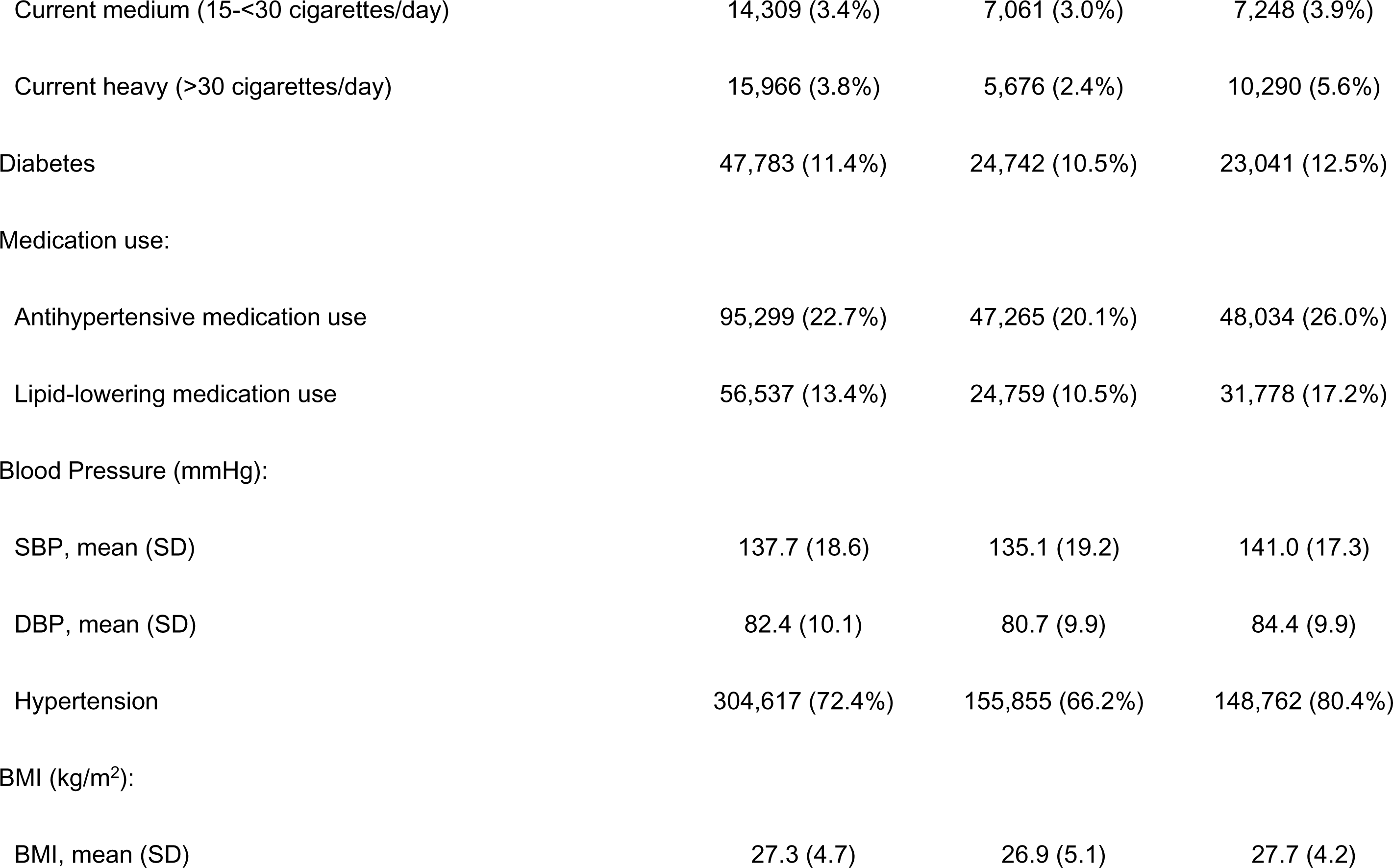
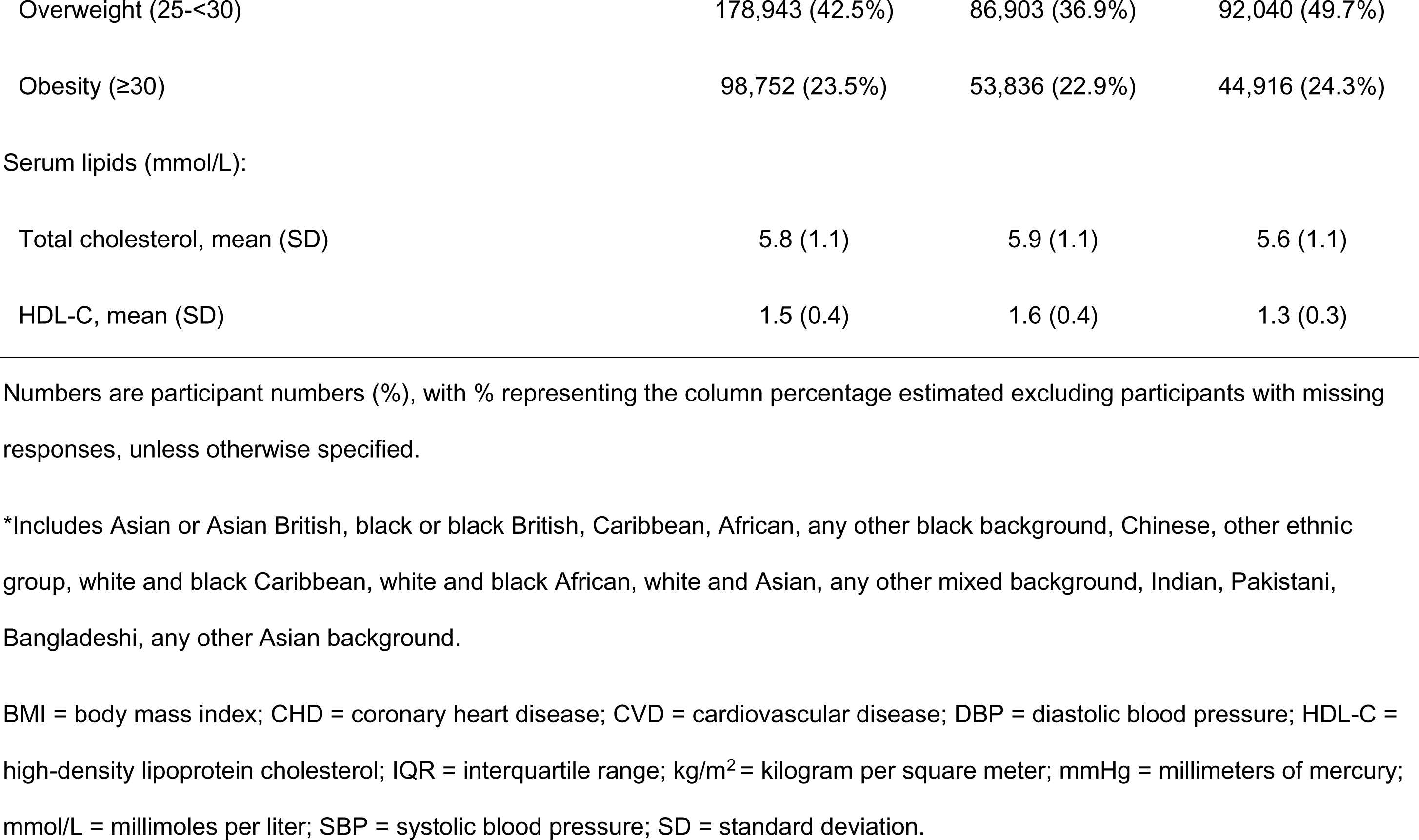
Baseline characteristics and incident cardiovascular disease events for 420,649 UK Biobank participants.

|  | Overall | Women | Men |
| --- | --- | --- | --- |
| Characteristics | (n = 420,649) | (n = 235,556) | (n = 185,093) |
| Incident total CVD cases during follow-up, n | 28,628 | 10,331 | 18,297 |
| Incident CHD cases during follow-up | 21,146 | 6,774 | 14,372 |
| Incident stroke cases during follow-up | 8,321 | 3,816 | 4,505 |
| Age, mean (SD) | 56.1 (8.1) | 56.1 (8.0) | 56.1 (8.2) |
| Ethnicity |  |  |  |
| White | 395,023 (94.4%) | 221,497 (94.4%) | 173,526 (94.3%) |
| Other* | 23,587 (5.6%) | 13,096 (5.6%) | 10,491 (5.7%) |
| Socioeconomic status |  |  |  |
| Townsend deprivation index score, median (IQR) | -2.2 (-3.7-0.4) | -2.2 (-3.6-0.4) | -2.2 (-3.7-0.5) |
| Townsend deprivation fifths: |  |  |  |
| First (least deprived) | 158,076 (37.6%) | 88,349 (37.5%) | 69,727 (37.7%) |
| Second | 86,275 (20.5%) | 48,820 (20.7%) | 37,455 (20.2%) |
| Third | 62,888 (15.0%) | 35,753 (15.2%) | 27,135 (14.7%) |
| Fourth | 56,033 (13.3%) | 31,621 (13.4%) | 24,412 (13.2%) |
| Fifth (most deprived) | 57,377 (13.6%) | 31,013 (13.2%) | 26,364 (14.2%) |
| Smoking status: |  |  |  |
| Never | 234,198 (56.0%) | 141,014 (60.2%) | 93,184 (50.6%) |
| Previous | 141,099 (33.7%) | 72,978 (31.1%) | 68,121 (37.0%) |
| Current light (<15 cigarettes/day) | 13,015 (3.1%) | 7,698 (3.3%) | 5,317 (2.9%) |
| Current medium (15-<30 cigarettes/day) | 14,309 (3.4%) | 7,061 (3.0%) | 7,248 (3.9%) |
| Current heavy (>30 cigarettes/day) | 15,966 (3.8%) | 5,676 (2.4%) | 10,290 (5.6%) |
| Diabetes | 47,783 (11.4%) | 24,742 (10.5%) | 23,041 (12.5%) |
| Medication use: |  |  |  |
| Antihypertensive medication use | 95,299 (22.7%) | 47,265 (20.1%) | 48,034 (26.0%) |
| Lipid-lowering medication use | 56,537 (13.4%) | 24,759 (10.5%) | 31,778 (17.2%) |
| Blood Pressure (mmHg): |  |  |  |
| SBP, mean (SD) | 137.7 (18.6) | 135.1 (19.2) | 141.0 (17.3) |
| DBP, mean (SD) | 82.4 (10.1) | 80.7 (9.9) | 84.4 (9.9) |
| Hypertension | 304,617 (72.4%) | 155,855 (66.2%) | 148,762 (80.4%) |
| BMI (kg/m<sup>2</sup>): |  |  |  |
| BMI, mean (SD) | 27.3 (4.7) | 26.9 (5.1) | 27.7 (4.2) |
| Overweight (25-<30) | 178,943 (42.5%) | 86,903 (36.9%) | 92,040 (49.7%) |
| Obesity (≥30) | 98,752 (23.5%) | 53,836 (22.9%) | 44,916 (24.3%) |
| Serum lipids (mmol/L): |  |  |  |
| Total cholesterol, mean (SD) | 5.8 (1.1) | 5.9 (1.1) | 5.6 (1.1) |
| HDL-C, mean (SD) | 1.5 (0.4) | 1.6 (0.4) | 1.3 (0.3) |

Numbers are participant numbers (%), with % representing the column percentage estimated excluding participants with missing responses, unless otherwise specified.
\*Includes Asian or Asian British, black or black British, Caribbean, African, any other black background, Chinese, other ethnic group, white and black Caribbean, white and black African, white and Asian, any other mixed background, Indian, Pakistani, Bangladeshi, any other Asian background.
BMI = body mass index; CHD = coronary heart disease; CVD = cardiovascular disease; DBP = diastolic blood pressure; HDL-C = high-density lipoprotein cholesterol; IQR = interquartile range; kg/m<sup>2</sup> = kilogram per square meter; mmHg = millimeters of mercury; mmol/L = millimoles per liter; SBP = systolic blood pressure; SD = standard deviation.

### Sex-specific risks and risk differences

During a mean follow-up of 13.2 years, there were 28,628 (10,331 women), 21,146 (6,774 women) and 8,321 (3,816 women) cases of incident CVD, CHD and stroke, respectively. Age-adjusted risks of incident CVD showed a “J-shaped” pattern and were higher among men in all categories of SBP and DBP (**Figure 1**). Risks of incident CVD were lowest at SBP 100-<105 mmHg for women (events per 10,000 person-years [95% CI], 15.6 [11.8-19.4]) and 110-<115 mmHg for men (47.2 [41.4-53.0]). Risks of incident CVD were lowest at DBP 60-<65 mmHg for women (22.7 [19.7-25.8]) and 65-<70 mmHg for men (63.3 [58.4-68.1]). Compared with SBP 100-<115 mmHg and DBP 70-<75 mmHg, sex-specific risk differences at higher levels of BP were larger among men than women (Table S5). We found similar patterns for CHD and stroke (Figures S2-S3 and Tables S6-S7).

**Figure 1.**
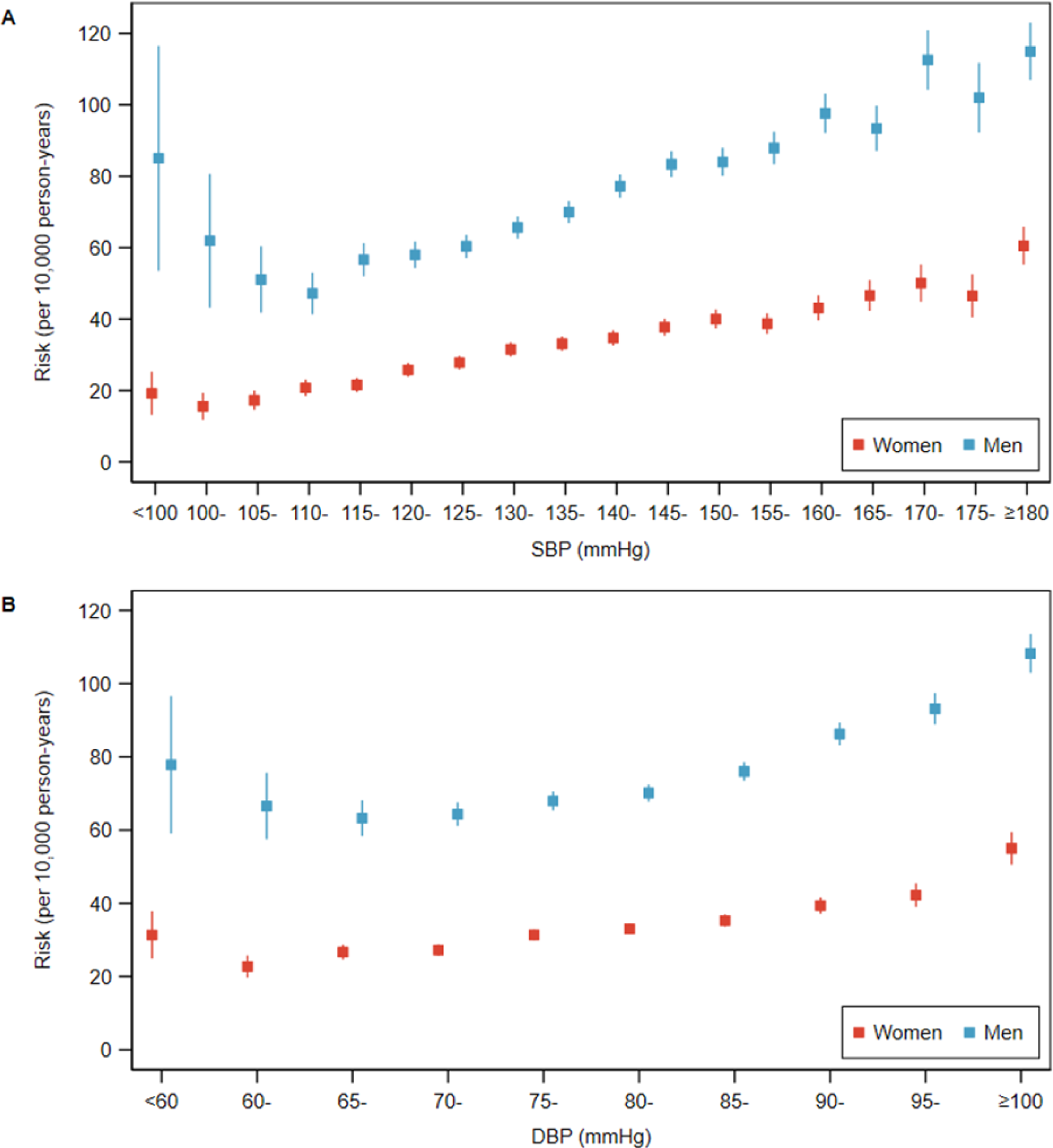
Risks of cardiovascular disease by level of blood pressure for each sex. Scatterplot showing age-adjusted risks (per 10,000 person-years) for incident cardiovascular disease across categories of (A) SBP and (B) DBP in women (red) and men (blue). Vertical lines indicate corresponding 95% confidence intervals. DBP = diastolic blood pressure; mmHg = millimeters of mercury; SBP = systolic blood pressure.

### Sex-specific relative risks

As would be expected from the risks, for SBP, there was an approximately “J-shaped” relationship for RRs of incident CVD for both sexes, which was roughly log-linear from nadirs of approximately 105 mmHg in women and 115 mmHg in men. Similarly, for DBP, with roughly log-linear increases from nadirs of approximately 65 mmHg in women and 75 mmHg in men (**Figure 2**). Sex-specific RRs for incident CVD, relative to 110-<115 mmHg for SBP and 70-<75 mmHg for DBP, were lowest at SBP 100-<105 mmHg (RR [95%CI], 0.76 [0.58-0.99]) and DBP 60-<65 mmHg (0.84 [0.72-0.97]) among women, and lowest at SBP 110-<115 (reference category) and DBP 65-<70 mmHg (0.98 [0.90-1.08]) for men, although CIs overlapped between adjacent BP groups. The association between higher SBP and increased RR was steeper among women compared with men. The patterns of sex differences in the associations of BP categories in relation to incident CHD and stroke were broadly similar, but with expected wider CIs (Figures S4-S5). The association between hypertension and CVD was stronger in women (RR hypertension vs no hypertension [95%CI], 1.62 [1.54-1.70]) than men (1.45 [1.39-1.52]) (Table S8). Stronger RRs for the associations between continuous BP and CVD were present among women than men. Equivalent, broadly similar, values for multivariable-adjusted RRs for the sex-specific associations of BP with all CVD outcomes are shown in Tables S8-S10.

**Figure 2.**
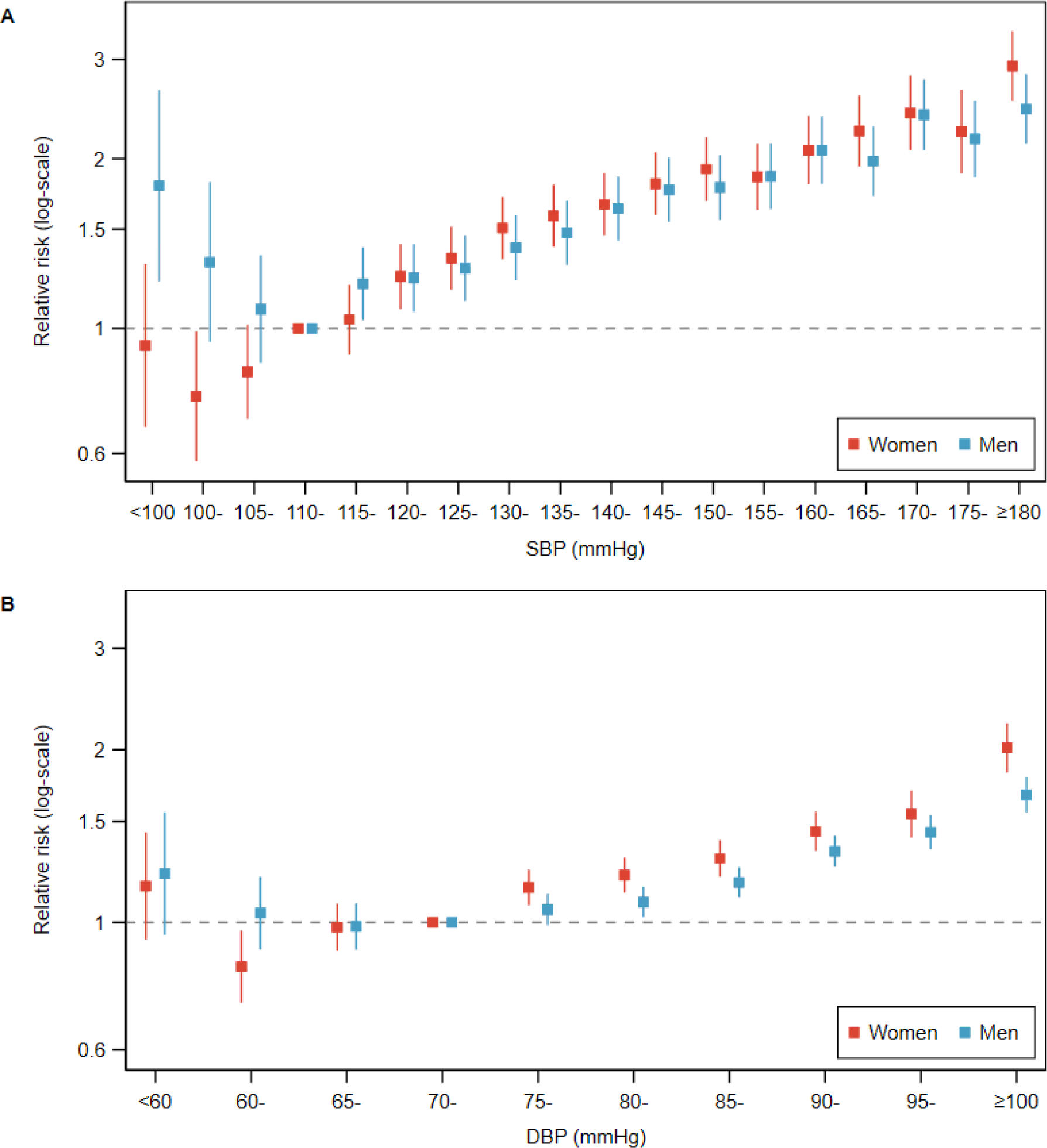
Sex-specific relative risks relating blood pressure to cardiovascular disease. Scatterplot showing age-adjusted sex-specific relative risks for incident cardiovascular disease across categories of (A) SBP and (B) DBP in women (red) and men (blue). Vertical lines indicate corresponding 95% confidence intervals. The reference category for SBP was 110-115 mmHg in both sexes separately. The reference category for DBP was 70-<75 mmHg in both sexes separately. DBP = diastolic blood pressure; mmHg = millimeters of mercury; SBP = systolic blood pressure.

### Sex-combined relative risks

When comparing both women and men to the same reference group, men with SBP 110-<115 mmHg (that is, men who had the lowest CVD risk), RRs of incident CVD in women were never higher than unity up to the 170<175 mmHg group and peaked at >180 mmHg when the RR was 1.32 (1.13, 1.53) (**Figure 3**). So, it is only women with SBP above 170 mmHg that had greater CVD risk than any man, in terms of risk according to SBP level. For DBP level, no category of women had higher risks than the lowest risk group for men of 70-<75 mmHg. Similar patterns were observed for CHD and stroke (Figures S6-S7). See Tables S11-S13 for age-adjusted and multivariable-adjusted sex-combined RRs for all CVD outcomes.

**Figure 3.**
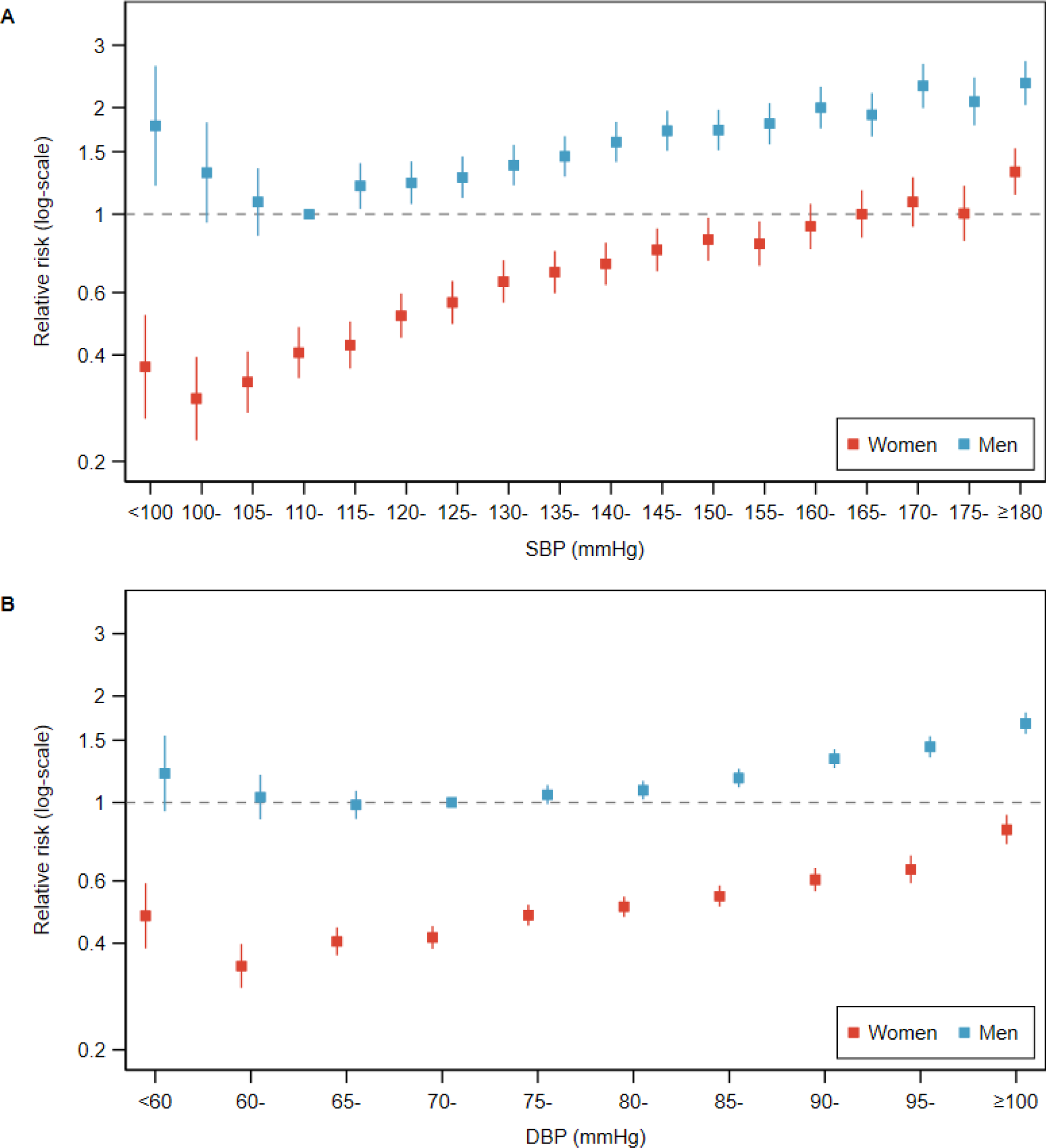
Sex-combined relative risks relating blood pressure to cardiovascular disease. Scatterplot showing age-adjusted sex-specific relative risks for incident cardiovascular disease across categories of (A) SBP and (B) DBP in women (red) and men (blue). Vertical lines indicate corresponding 95% confidence intervals. The reference category for SBP was men at 110-115 mmHg for both women and men. The reference category for DBP was men at 70-<75 mmHg for both women and men. DBP = diastolic blood pressure; mmHg = millimeters of mercury; SBP = systolic blood pressure.

### Subgroup analyses

The patterns of BP-CVD relationships were much the same in subgroup analyses by age (**Figure 4**, Table S14) and by menopause status in women (Table S15). RRs were greater for the associations between BP and incident CVD among younger age groups for both sexes and for pre-menopausal women compared with menopausal women, presumably due to the increasing risk with age.

**Figure 4.**
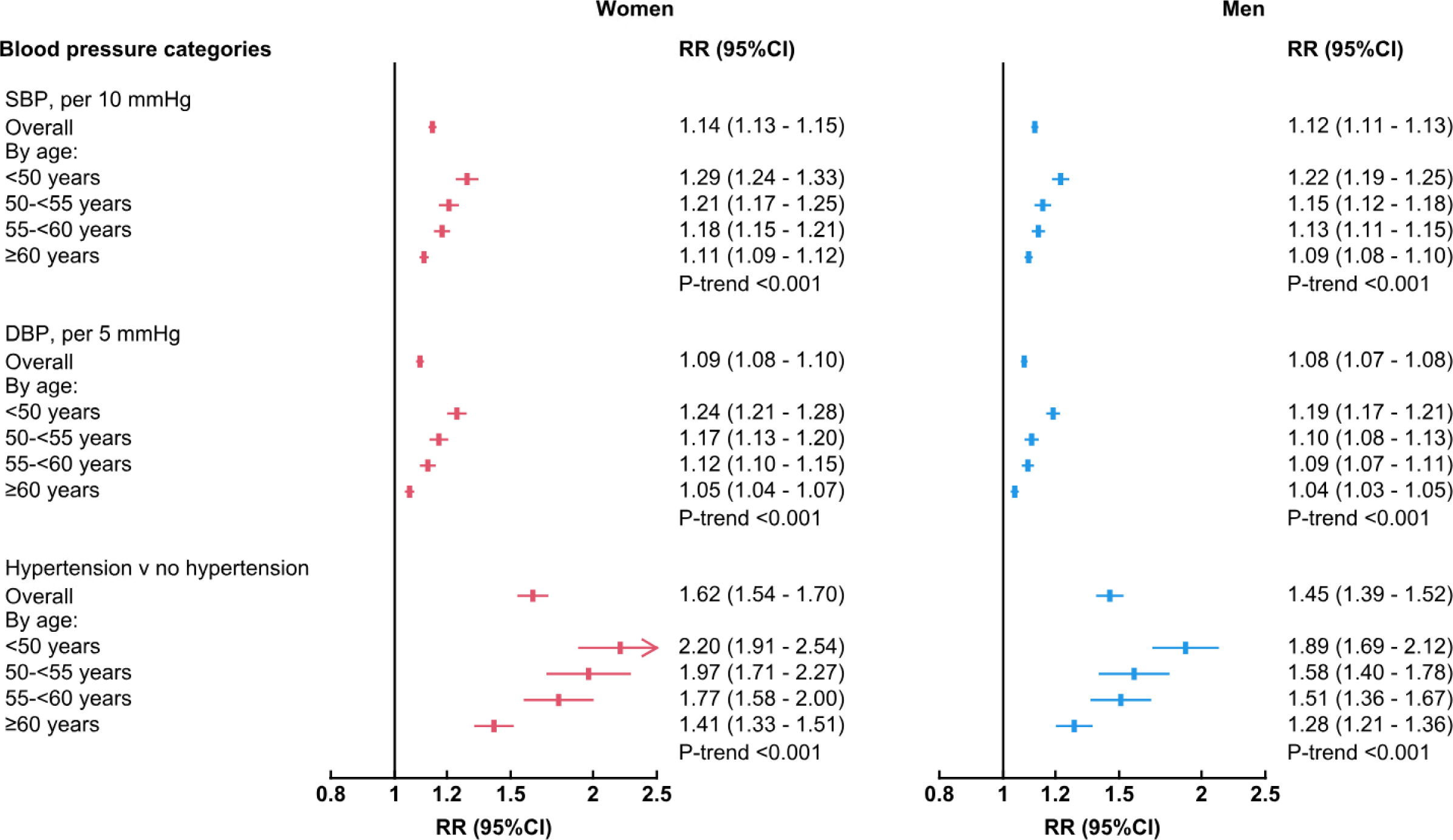
Sex-specific relative risks relating blood pressure level to cardiovascular disease by sex and age group. Forest plots showing sex and age-specific RRs relating continuous blood pressure and hypertension status to incident cardiovascular disease in women (red) and men (blue). Horizontal lines indicate corresponding 95% confidence intervals. CI = confidence interval; DBP = diastolic blood pressure; mmHg = millimeters of mercury; SBP = systolic blood pressure; RR = relative risk.

### Sensitivity analyses

When excluding participants taking antihypertensive medication at baseline, the directions of association remained similar, although the magnitude of RRs for CVD generally became stronger (Table S16).

Among 51,375 participants with baseline and follow-up BP measurements, regression dilution ratios for SBP and DBP were estimated to be 0.624 and 0.536, respectively, for women and 0.551 and 0.455, respectively, for men (Figure S8-S9). Baseline characteristics were similar between participants with baseline and follow-up BP measurements and participants in the main study sample (Table S17). After correction for regression dilution, the sex-specific age-adjusted RRs by SBP and DBP levels for women moved slightly closer to those for men (Figure S10).

## Discussion

This large study assessed sex-differences in the associations of BP with CVD, as well as CHD and stroke subtypes. CVD risks were higher among men than women at all levels of BP. Whilst there was a clear indication of a log-linear association starting substantially lower than current AHA treatment thresholds of SBP >130 mmHg and DBP >80 mmHg for both sexes. There was a “J-shaped” pattern relating increasing BP to CVD outcomes in both sexes, with a lower nadir in women (SBP 100-<105 mmHg and DBP 60-<65 mmHg) than men (SBP 110->115 mmHg and DBP mmHg). RRs comparing to the reference groups of SBP 110-<115 or DBP 70-<75 were mostly higher for women than for men when the sexes were analyzed totally separately, but RDs showed the opposite relationship (**Figure 5**). Furthermore, when a common reference group of men with SBP at 110-115 mmHg or men at DBP 70-<75 mmHg was used, when analyzing the sexes together, the RRs for women were lower than for men at all BP levels, whilst the risk of CVD in women only exceeded that of men at their lowest level of risk at high levels of BP.

**Figure 5.**
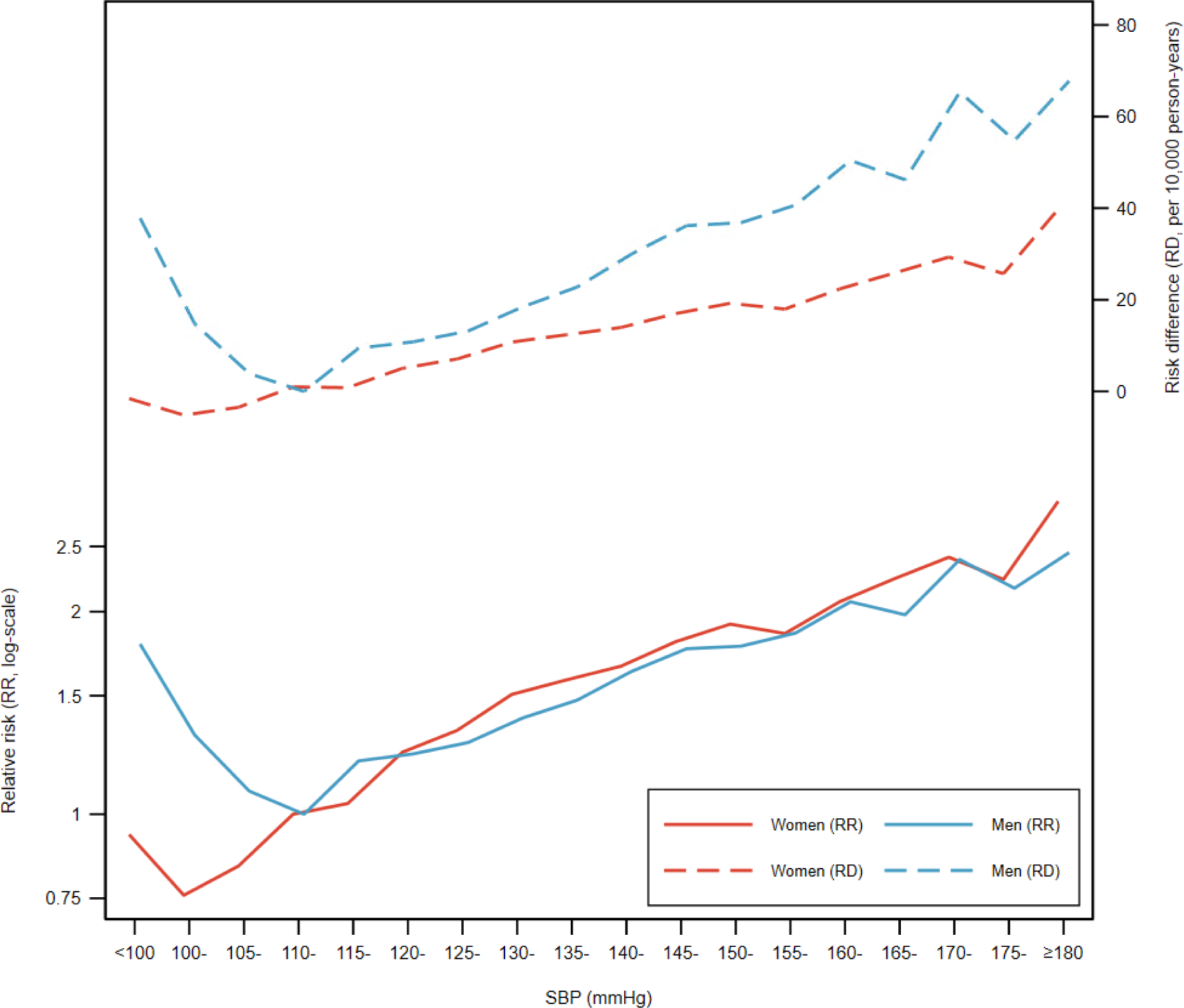
Sex-specific risk differences and relative risks relating systolic blood pressure to incident cardiovascular disease. Line graph showing age-adjusted sex-specific RDs per 10,000 person-years (dashed lines) and RRs (solid lines) for incident cardiovascular disease across categories of SBP in women (red) and men (blue). The reference category for SBP was 110-115 mmHg in both sexes separately. DBP = diastolic blood pressure; mmHg = millimeters of mercury; RD = risk difference; RR = relative risk; SBP = systolic blood pressure.

There is established evidence that exceeding the existing thresholds for normal BP (SBP > 120 mmHg or DBP >80 mmHg) is associated with an elevated risk of CVD in both sexes.^22–24^ However, few studies have explored associations of BP levels below these thresholds with CVD risks, or considered BP categories as narrow as in this study.^4,8,25^ A recent study of 53,289 participants in the National Health and Nutrition Examination Survey (NHANES) found the risk of CVD mortality was lowest at SBP 100-<110 mmHg in women and 110-<120 mmHg in men.^8^ Another study of 27,452 participants in four community-based cohorts found risks of CVD events were lowest at SBP <100 mmHg in women and <130 mmHg in men.^4^ Other studies have considered CHD and stroke subtypes and, as in the current study, similar patterns were reported to those with CVD ^4,6^. In this study there were higher risks present at around SBP <100 mmHg and DBP <60 mmHg, compared to slightly higher BP levels, for both sexes. This appeared to be more pronounced for SBP among men, as well as among women who were menopausal or ≥55 years of age in subgroup analyses. Investigators analyzing NHANES data identified the same pattern for SBP and CVD mortality for both men and women.^8^ One explanation is that lower SBP is associated with higher mortality among patients with heart failure ^26^ and older adults (≥80) with frailty,^27^ although our study excluded those with pre-existing CVD and was a predominantly middle-aged cohort.

Prior epidemiological studies suggest that the strength of associations between BP and CVD varies by sex.^4,5,7,8,28,29^ Meta-analyses of observational studies have found that RRs for the association of BP with CVD events ^12^ and mortality ^11,30^ are stronger in women than men, although all studies concluded that these sex-based differences were not substantial. The most recent meta-analysis of 11 observational studies reported that AHA stage 1 hypertension was associated with 50% and 37% higher risks of CVD compared to normal BP in women and men, respectively.^12^ Similarly, the present study found that hypertension was associated with a higher excess risk for incident CVD in women (46% higher) than men (32% higher) compared to no hypertension. However, CVD risks remained higher in men than women for hypertension and across all levels of BP.

Studies consistently demonstrate that women have lower mean basal BP than men.^31,32^ The trajectory of BP may differ by sex over time, with women exhibiting faster rise in BP from as early as the third decade of life.^33^ The present study examined the roles of age and menopausal status, finding higher BP was associated with larger RRs for CVD among women that were pre-menopausal or younger (<55 years). The influence of age on the relationship between BP and CVD was more pronounced among women than men, with differences between age, sex and BP observed for all BP metrics.

Hormones, as well as anatomic and physiological differences, may explain sex-differences in optimal BP and BP trajectories over time.^9^ Specifically, estrogen may exert benefit effects on the renin-angiotensin-aldosterone system influencing BP homeostasis, and increased plasma renin and sympathetic nerve activity and decreased nitric oxide have been observed in post-menopausal women.^34–37^ Compared with men, women have smaller coronary arteries, smaller aortic root dimensions and lower aortic distensibility with age, as well as higher pulse pressure, which may make women more susceptible to the cardiovascular effects of hypertension.^38–40^ Moreover some conditions specific to women, including premature menopause and pregnancy-associated hypertension, may worsen CVD outcomes when coincident with hypertension.^41–43^ Lastly, emerging evidence suggests that the effect of genetic variants for BP are greater in women than men.^44^

Treatment for high BP is typically through antihypertensive drug therapy, although lifestyle changes, including salt reduction, can be expected to assist. In questioning whether women should be treated differently to men it is necessary to consider whether they react to the drugs differently. Individual clinical trials have described sex-differences in the efficacy, pharmacodynamics and pharmacokinetics, and overall women report more adverse effects from antihypertensive medication.(6,7) However, the BP Lowering Treatment Trialists’ Collaboration has found that, with antihypertensive drug treatment, BP reductions were similar for women and men in every comparison they made, and there was no sex difference in protection against CVD regardless of age or antihypertensive regimen.^13,30^

### Study strengths and limitations

Strengths of this study include the prospective study design and large sample size, which allowed us to examine smaller ranges of BP than previously studied, as well as major CVD subtypes. We also considered the roles of age and menopausal status in more detail than prior studies. There are limitations to our study. BP measurements were taken at a single visit, which may be prone to measurement error.^2^ Indeed, our assessment of regression dilution bias in BP using participants with repeat measurements suggests this may have led to an underestimation of reported RRs, particularly among men.^45^ Because of the extensive baseline data available in UK Biobank, we were able to account for potential confounders, including serum total cholesterol and HDL-C. However, this study was unable to account for changes in these factors during the follow-up period, and we cannot exclude the effects of unmeasured and residual confounding, particularly in antihypertensive medication use. Reverse causation also cannot be ruled out, although we excluded participants with prior CVD at baseline. Finally, the UK Biobank study population is not nationally representative,^46^ being predominantly of white ethnicity and higher than average social advantage, and so our findings may be prone to selection bias, particularly for risk estimation, although risk factor associations still appear to be generalizable.^47^ Lastly, we only studied CVD, whereas normal or optimal BP for other disease outcomes may differ, and the potential adverse effects at lower BP levels were not examined.

### Conclusions

In this study we have separated the effects of BP levels by risk and RR, which have been confused in past literature (1) and considered narrow increments of BP. Our analyses suggest that the level at which hypertension is defined should not vary by sex. As regards to determining the level at which antihypertensive medication should be initiated to prevent CVD, the definition of hypertension itself is moot, since intervention should depend upon risk, not BP level alone,^48,49^ provided that risk has been estimated using a sex-specific algorithm.

## Data Availability

Bona fide researchers can apply to use the UK Biobank dataset by registering and applying at http://ukbiobank.ac.uk/register-apply/.

## Acknowledgements

This research has been conducted using the UK Biobank Resource under application number 74018. We thank all participants, researchers and support staff who make the study possible. Bona fide researchers can apply to use the UK Biobank dataset by registering and applying at http://ukbiobank.ac.uk/register-apply/.

## Supplemental Material

Figures S1-S10

Tables S1-S17

## Non-standard Abbreviations and Acronyms

ACC: American College of Cardiology
AHA: American Heart Association
LRT: likelihood ratio test
NHANES: National Health and Nutrition Examination Survey
OPCS: Operating Procedure Codes Supplement
RD: risk difference

